# Gender- and age-related differences in misuse of face masks in COVID-19 prevention in central European cities

**DOI:** 10.1101/2020.11.11.20224030

**Authors:** Linda Eckl, Stefan Hansch

## Abstract

1

**Objective:** Correct use of face masks is required for their efficacy in preventing possible droplet infections with SARS-CoV-2. We tried to provide information about differences in the distribution of gender and age groups wearing face masks incorrectly.

**Design:** Pilot field study

**Methods:** Visual observation of mask use in public, not age- and gender-related places in central European large cities regarding incorrect mask-wearing (n=523); statistical analysis (nominal scale) in terms of gender and estimated age group using the total numbers, binomial test and chi-square test.

**Results:** There is no significant difference (binomial test: p-value = 0.43) in mask misuse between the genders (female: 271 (51.8%), male: 252 (48.2%) and 0 non-binary individuals (0%)). There is a significant difference (chi-square test: p-value < 2.2e-16) in age group distribution (170 young 10-29 years (32.5%), 261 middle-aged 30-59 years (49.9%), 92 older adults ≥ 60 years (17.6%)). In total numbers, the highest counts were observed in middle-aged persons with 261 counts (49.9%).

**Conclusion:** Our study shows an uneven age-distribution of people wearing the face mask in public improperly.

## 2 INTRODUCTION

In the spread of the global pandemic of corona virus disease 2019 (COVID-19) caused by the severe acute respiratory syndrome coronavirus 2 (SARS-CoV-2), the necessity of protective countermeasures emerges. Dependent upon geographical area, professional or community setting, the use of face masks is controversially discussed or often mandatory.

A recent overview by Feng et al.[1] shows how different healthcare authorities handle the current evidence by providing different recommendations on the use of face masks. Despite initial discouragement or at least insufficient evidence of risk reduction to get infected, the wearing of face masks in defined situations was recommended by the German Federal Ministry of Health[2]. Public guidelines for the use of face masks were provided by the German Federal Institute for Drugs and Medical Devices[3], inter alia regarding the placement of a closely fitting mask covering mouth and nose.

Besides other measures like distance and hand hygiene, the optimal use of face masks is needed to provide a sufficient efficacy of the physical barrier’s aspired protective effect[4]. A recent study by Leung et al.[5] on expiratory virus shedding found higher virus loads in nasal swabs than in throat swabs, so an incorrectly worn mask without covering the nose could drastically facilitate disease spread.

The prevalence of masking after the post-lockdown reopening of businesses has currently been examined in other studies[6, 7], and significant differences in regional, age and gender groups have been found.

We address the question: who are the people in public that are not wearing the face mask properly and therefore risk to spread SARS-CoV-2? This could contribute to improve health behavior education campaigns and / or advertisements, under the assumption, although not proven by this study, that people, who are wearing a mask, and therefore already show compliance with the latest regulations, are generally approachable for the correct use of a mask and prevention measurements. Three factors influenced the observations: I) the general age- and gender-distribution in public, II) the probability that a certain age- and gender-group does not properly use the mask and III) the distribution seen by the observers.

For efficiency reasons with focus on the spreading of SARS-CoV-2 in public space, we combined the numbers of the general age- and gender-distribution in public (I) and the probability, a certain age- and gender-group does not properly use the mask (II) as there is lack of consequences in the first line, if surveyed separately. Combining these two factors leads to a distribution of people improperly using the face masks in the public, which itself could risk a higher spread of the infections. Factor III was tried to be minimized, as to be seen in the Methods part of this manuscript.

We therefore measured the total counts of different age- and gender-groups in public (I and II combined), that are using the mask incorrectly.

## 3 METHODS

Between June 2020 and September 2020, we performed observations at different times and weekdays in three German (Regensburg, Augsburg and Berlin), one Austrian (Vienna), and one Polish city (Szczecin). These observations were conducted by a team of one female and one male researcher and took place in public places and transportation (buses, streetcars, subways, trains, stations, shopping malls, bakeries and supermarkets). Each of these places had an official recommendation to wear a face mask. We excluded gender- and age-specific locations, like woman clothing-shops, schools or retirement homes. The male and the female researcher both were medically experienced in signs of pre-aging and physical signs of age. Both researchers had to see the person, otherwise the person was excluded. Furthermore, both had to confirm the age-group and gender. Without confirmation of both scientists, physical signs, like specific clothing, physical signs or family-status were used to determine an age group.

As an incorrect fit of face mask, we defined any deviant kinds of mask use, i.e., covering only the mouth or the nose but not both or neither of them, a very loose fit with forming gaps between the mask and the face and taking the mask off for coughing or sneezing.

As correct use of the face mask served the WHO recommendation, as to be seen in the “Advice on the use of masks in the context of COVID-19”[8].

We considered all people in the mentioned public places as eligible, with the exclusion criteria of (1) children under the estimated age of 10 years and (2) individuals with visible physical disabilities like people in wheelchairs, with walking aid or oxygen device.

Age groups were divided into young (10-29 years), middle- (30-59 years), and advanced age (≥ 60 years), independently estimated by the two researchers. Gender groups included phenotypically female, male, and non-binary gender. Observations were taken from a distance without interaction between researchers and subjects.

Notes were taken digitally by a tally list in the above-mentioned categories. We conducted statistical analysis (nominal scale) in terms of gender and age, total numbers, binomial test, and chi-square test using the R-functions chisq.test() and binom.test().

We tried to further minimize the above-mentioned distribution seen by the observers (III) by different approaches. One of these approaches concerns the time of the day. We performed separate measurements, either between morning and noon (9 am – 2 pm, three measurements) or in the afternoon (4 pm - 8 pm, three measurements), for an average duration of 1.5 hours for each observation. Another approach was made by watching different cities. Both are analyzed as part of the whole statistical analyses and separately.

There was no acquiring of personal data, so no conclusions on individuals can be drawn. Therefore, the University of Regensburg ethics committee saw a board review of our study as not obligatory.

## 4 RESULTS

We observed 523 samples of incorrect mask use, shown in Table 1 and Figure 1.

**Table 1:** Number and percentages of counts per gender and age group.

| | Young<br>(10-29 years) | Middle-aged<br>(30-59 years) | Advanced-aged ($\geq 60$<br>years) | total |
| --- | --- | --- | --- | --- |
| male | 80 (15.3%) | 131 (25.0%) | 41 (7.8%) | 252 (48.2%) |
| female | 90 (17.2%) | 130 (24.9%) | 51 (9.8%) | 271 (51.8%) |
| total | 170 (32.5%) | 261 (49.9%) | 92 (17.6%) | 523 (100%) |

**Figure 1.**
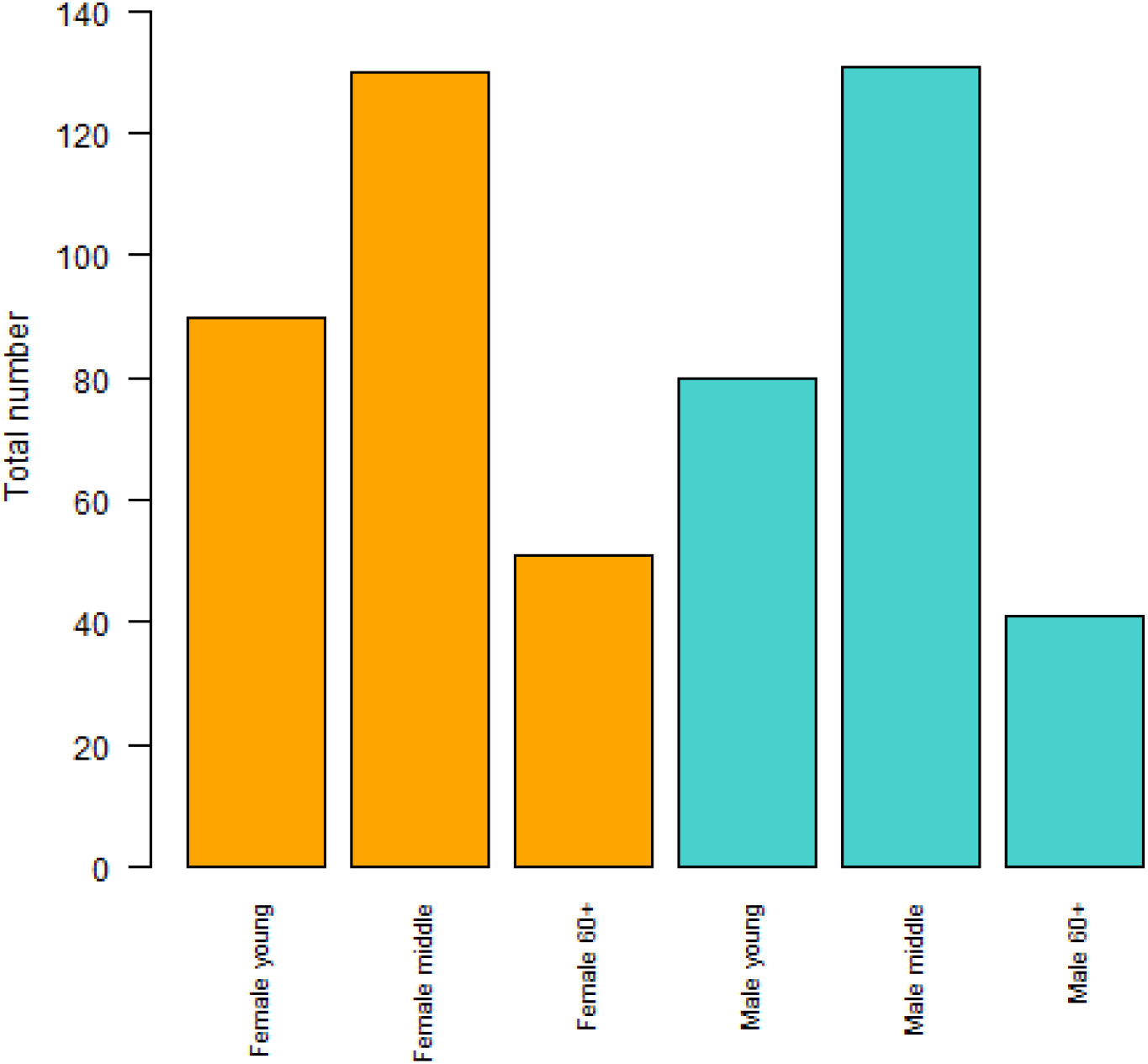
Total numbers of observed subjects.

The first question was if there is a difference in misuse of the face mask between the genders. As we observed no cases in the non-binary gender group, only male and female genders are mentioned in the following.

Our hypothesis was, that there is no difference between man and woman in misuse. Therefore, we compared the total numbers of each gender (271 (female) vs. 252 (male)) using the binomial test and received a p-value = 0.4313. This p-value is above our significance level of p<0.05. Furthermore, we applied the chi-square test on our data, which showed *X-squared = 0*.*99008, df = 2, p-value = 0*.*6095*, which also indicates no difference.

Another question was if there is a difference between the age-groups (young with 170, middle-aged with 261 and older adults with 92 persons). We also applied a chi-square test and received *X-squared = 82*.*076, df = 2, p-value < 2*.*2e-16*. This p-value is below p<0.05 and shows, that misuse of face masks is significantly age-related. In total numbers, the highest counts were observed in middle-aged persons with 261 counts (49.9%). Further analyses, which are especially addressing different cities and different daytimes are shown in Table 2 and Table 3. All these analyses are statistically significant.

**Table 2:** We analyzed if there is a difference in the age-distribution between Regensburg (most data) and four other cities. Three observations were taken between morning and noon (9 am - 2 pm), and three observations were taken in the afternoon (4 pm - 8 pm). The measurements typically lasted for 1.5 hours. With a significance level below p<0.05, the data above show an uneven age-distribution calculated with chi-square test.

|  | Young | Middle-aged | Advanced-aged | Qui-square p-value |
| --- | --- | --- | --- | --- |
| Outside Regensburg | 75 | 127 | 50 | 1.053e-08 |
| Regensburg | 95 | 134 | 42 | 5.603e-11 |
| total | 170 | 261 | 92 | 2.2e-16 |
| Morning + Noon | 24 | 58 | 23 | 1.185e-05 |
| Afternoon | 51 | 54 | 10 | 1.423e-07 |

**Table 3:** We analyzed if there is a difference in the gender groups between Regensburg (most data) and four other cities. Three observations were taken between morning and noon (9 am - 2 pm), and three observations were taken in the afternoon (4 pm - 8 pm). The measurements typically lasted for 1.5 hours. With a significance level above p>0.05, the data above show no difference between the genders in misuse of the masks, p was calculated with the binomial test.

|  | Female | Male | Binomial p-value |
| --- | --- | --- | --- |
| Outside Regensburg | 140 | 112 | 0.08877 |
| Regensburg | 131 | 140 | 0.6271 |
| total | 271 | 252 | 0.4313 |
| Morning + Noon | 53 | 52 | 1 |
| Afternoon | 60 | 55 | 0.7093 |

## 5 DISCUSSION

Our study aims to determine if there is a relevant difference in the misuse of face masks between subgroups in terms of gender and age.

This study is the first one to assess the misuse of face masks during the current global pandemic. This topic is especially of interest, to evaluate the application of protective countermeasures in public, and it can contribute to further modelling and prognosis of pandemic spread.

Another strength is the relative high case number, which allows a representative statement about our study question, and makes it comparable to other works. We performed our observations in cities in the south (Regensburg, Augsburg) and the northeast of Germany (Berlin), in a large Polish city (Szczecin), and the capital city of Austria (Vienna) to improve the study design by decreasing regional effects. Here must be said that the highest number of observations was taken in Regensburg, but a cross-analysis (see tables 2 and 3) between Regensburg and the other cities shows a significantly uneven distribution of age groups with p-values below our level of significance of <0.05.

Also, some limitations must be noted. One is the time window of our observations in terms of the whole timespan. There may be an effect of in- or decrease of mask misuse over time in the assessed places, influenced by the current climate of public opinion or the continuously updated state of knowledge.

Another limitation lies within the location of our observations. These were taken mainly in centrally located malls, shops, supermarkets and stations of large cities as described in methods. Rural areas, senior homes, universities, and suburban regions are not included in our study. Further and more widespread investigation can contribute to exploring the topic of mask misuse in these areas.

## 6 CONCLUSION AND FUTURE WORK

Our pilot-study shows a significant, gender-independent difference between age groups in the correct use of face masks in public. This serves as a reference point for our further investigation concerning subgroup-analyses and prevalence studies, which are already in preparation. The results may be useful in health education and advertising, like promoting to avoid public places or the correct mask use, addressing especially subgroups with highest misuse rates.

## Supporting information

EQUATOR-Checklist

## Data Availability

The data that supports the findings of the study are available from the corresponding author, S.H., upon reasonable request

## 8 ACKNOWLEDGMENTS

We thank Florian Schlieckau, Simon Stelzl, Karl-Peter Ittner, Gerhard Hansch, and Fro Wirtz for helpful discussions

